# Measuring Hemoglobin A1C with Frozen Packed Cell and Frozen Whole Blood Samples in an Epidemiologic Study: The Reasons for Geographic and Racial Differences in Stroke (REGARDS) Study

**DOI:** 10.1101/2023.09.27.23296242

**Authors:** Debora Kamin Mukaz, Stephanie Tison, D. Leann Long, April P. Carson, Kelly J. Hunt, Suzanne E. Judd, Mary Cushman

## Abstract

**Introduction:** Hemoglobin A1c (HbA1c) measurement in epidemiology studies could be increased if reliability of measurements in frozen stored samples was known. In the REasons for Geographic and Racial Differences in Stroke, a longitudinal study of 30,239 Black and White U.S. adults, we investigated reliability of HbA1c measurements for two types of samples stored at -80°C for up to 14 years.

**Methods:** Among 917 participants without diabetes, HbA1c was measured in 2017 in frozen packed cells from the first visit (2003-07) and in frozen whole blood samples from the second visit (2013-16). To study reliability, associations between HbA1c and glycemia-related characteristics were examined.

**Results:** Each 10mg/dL greater fasting glucose was associated with 0.08% (95%CI: 0.05-0.11%) greater HbA1c in frozen packed cells (visit 1) and 0.10% (95%CI: 0.08-0.12%) greater HbA1c in whole blood (visit 2). HbA1c was also similarly higher with both methods with increasing age, gender, systolic blood pressure, body mass index, high-density lipoprotein, triglycerides, C-reactive protein, and hemoglobin. Using both methods, ≤3.5% were classified with diabetes based on HbA1c ≥6.5%.

**Conclusions:** In REGARDS participants without diabetes, HbA1c measurement appeared reliable in frozen packed cells or whole blood under long-term storage, suggesting acceptability for study of the epidemiology of HbA1c.

## INTRODUCTION

Hemoglobin A1c (HbA1c) has long been used to assess glycemic control in the management of diabetes, and HbA1c level correlates with the risk of complications.^1^ With advances in assay standardization, use of HbA1c testing was incorporated into the diagnostic criteria for diabetes in 2010.^2^ HbA1c testing is typically performed on fresh whole blood samples, with samples refrigerated at 4°C stable for up to seven days. Prior studies, most which employed high-performance liquid chromatography (HPLC) assays for HbA1c, demonstrated the feasibility of testing frozen whole blood samples stored for months or years.^3-10^

The Reasons for Geographic and Racial Differences in Stroke (REGARDS) study is a longitudinal epidemiologic study of 30,239 Black and White adults aged 45 or over undertaken to ascertain the causes of the excess stroke mortality among Black people and those living in the Southeastern United States.^11^ Recruitment took place between 2003 and 2007, prior to use of HbA1c for diabetes classification, so fasting plasma glucose was the glycemic measure used in both the initial exam and the subsequent study exam in 2013 - 2016. Given the central role HbA1c testing now plays in diagnosis and treatment of diabetes and to study it as a risk factor and outcome, the REGARDS investigators desired adding its measurement to the study data. Immunoturbidimetric assays (immunoassays) are increasingly employed in clinical HbA1c testing because they are based on structural differences, not charge differences (as in ion-exchange HPLC) between glycated and non-glycated hemoglobin. Thus, the effects of the most common variant hemoglobins (e.g., hemoglobin S) on HbA1c measurement using immunoassays are mitigated.^12^

The purpose of this study was to assess whether REGARDS could provide measurement of HbA1c sufficiently accurate for study of the epidemiology of this biomarker. HbA1c was measured in a sample of REGARDS participants who were free of diabetes at both visits to determine whether values obtained by immunoassay of whole blood or packed cell samples stored at -80°C for up to 14 years were correlated with: a) results of fasting plasma glucose testing performed soon after sample collection, and b) glycemia-associated phenotypic characteristics.

## MATERIALS AND METHODS

### Study population and sample collection

#### Pilot Study

was performed to direct the plan for HbA1c method selection and assure we would likely obtain results sufficiently accurate for epidemiologic research. This included 27 adults aged 30 and older. Blood was collected in EDTA tubes. Non-centrifuged whole blood samples and packed cells were frozen at −80°C for 24-48 hours. Conduct of the pilot study was approved by the University of Vermont Institutional Review Board.

#### REGARDS Study

Between January 2003 and October 2007, REGARDS recruited 30,239 Black and White participants, aged 45 years and older, in the contiguous 48 states. The study oversampled Black individuals and residents of the stroke belt region in the southeastern United States (North Carolina, South Carolina, Georgia, Alabama, Mississippi, Tennessee, Arkansas, and Louisiana). Computer-assisted telephone interviews were used to obtain verbal informed consent, demographic information, and medical history. Written informed consent was obtained, and physiologic measurements and fasting blood samples were collected during an in-home visit, three to four weeks after the telephone interview. A detailed description of the study design has been described elsewhere.^11^ Between 2013 and 2016, a second in-home visit collected similar information using the same methods as the first visit. The protocol was reviewed and approved by Institutional Review Boards of participating institutions and all participants provided written informed consent.

At the first REGARDS visit, fasting morning blood was collected in EDTA plasma tubes, then stored on ice packs (0 °C) for the remainder of the visit.^13^ Centrifugation for 10 minutes was performed within two hours of phlebotomy, then serum or plasma, and packed cells from EDTA tubes were transferred into mailer tubes. The mailer tubes were shipped overnight with frozen gel ice packs to the University of Vermont’s Laboratory for Clinical Biochemistry Research (LCBR), where they were inventoried, re-centrifuged at 30,000xG (−4°C) and serum, plasma and the cell layer stored at −80°C.^13^ At the second visit the blood collection protocol was similar but added an EDTA whole blood sample that was not centrifuged, and which was stored at −80°C at the LCBR.

### Assay Methods

#### Pilot study

We compared HbA1c measurements from three sample types: fresh whole blood, frozen whole blood, and frozen packed cells (neat and 1:2 dilution in NaCl). Because many prior studies on HbA1c testing of frozen samples employed HPLC assays, testing of fresh whole blood samples was performed using an ion-exchange HPLC method (Tosoh G8, Tosoh Bioscience, South San Francisco, CA). Testing of frozen samples (whole blood and packed cells) was performed using the Cobas Integra 400 system (Roche Diagnostics, Indianapolis, IN).

#### REGARDS Study

HbA1c was measured in frozen packed cells from visit 1 and frozen whole blood from visit 2 (both from EDTA tubes) using an immunoturbidimetric method for hemolyzed whole blood (Tina-quant® HbA1c Gen. 3, Cobas Integra 400, Roche Diagnostics, Indianapolis, IN). All hemoglobin variants, which were glycated at the N-terminus of the β-chain and which had antibody-recognizable regions similar to HbA1c, were measured with the assay. Within-run coefficients of variation (CVs) were 1.0-1.6% and between-run CVs were 1.4-2.0%. For both visits, HbA1c was measured in 2017.

### Definition and Measurement of Glycemia-associated characteristics

Age was self-reported at both visits. Gender, race (Black and White), education and income were self-reported at the first visit only. Education was grouped into <high school, high school graduate, some college, and college graduate and above. Income was categorized as <$20k, $20k-$34k, $35k-$74k, $75k and above, or not willing to report. Self-reported smoking status was categorized into current, former, and never.

At both visits, blood pressure was measured twice using a sphygmomanometer after the participant rested for 5 minutes in a seated position; the two values were averaged. Body mass index (BMI) was calculated by dividing weight in kilograms by the square of the height in meters.

At the first visit, glucose was measured using colorimetric reflectance spectrophotometry on the Ortho Vitros 950 IRC Clinical Analyzer (Johnson & Johnson Clinical Diagnostics, Rochester, NY); 87 percent of participants adhered to the overnight fasting request.^14^ At the second visit, due to requirements for reporting results to participants, among participants residing in New York state, glucose was measured using the same method as the first visit. For other participants the Cobas Integra 400/800 system (Roche Diagnostics, Indianapolis, IN) was used. Diabetes status was defined as fasting glucose ≥126 mg/dL or random glucose ≥200 mg/dL or self-reported diabetes or use of oral medications or insulin.

At the first visit, High-density lipoprotein (HDL) cholesterol and triglycerides were measured in serum using the Ortho Vitros Clinical Chemistry System 950IRC instrument (Johnson & Johnson Clinical Diagnostics, Rochester, NY). For HDL cholesterol, samples were treated with dextran sulfate/magnesium chloride to precipitate chylomicrons, very-low-density lipoprotein (VLDL) cholesterol, and low-density lipoprotein (LDL) cholesterol. LDL cholesterol was calculated using the Friedwald equation (total Cholesterol – HDL cholesterol – (triglycerides/5).^15^ At the second visit, HDL cholesterol and triglycerides were measured using the Roche Cobas Integra 400/800 system (Roche Diagnostics, Indianopolis, IN).

At the first visit, serum creatinine was measured by colorimetric reflectance spectrophotometry on the Vitros 950IRC instrument (Johnson & Johnson Clinical Diagnostics, Rochester, NY) and creatinine was calibrated to isotope dilution mass spectrometry methods.^16^ Urinary albumin was measured using the BN ProSpec nephelometer (Dade Behring, Deerfield, IL now Siemens AG, Marburg, Germany) and urinary creatinine using the Modular-P chemistry analyzer (Roche/Hitachi, Indianapolis, Switzerland). Urine albumin-to-creatinine ratio (ACR) was calculated. Estimated glomerular filtration rate (eGFR) was calculated using the Chronic Kidney Disease Epidemiology Collaboration (CKD-EPI) equation.^17^ At the second visit, among participants residing in New York state, serum creatinine was measured using the same method as the first visit. For other participants, the Cobas Integra 400/800 system (Roche Diagnostics, Indianapolis, IN) was used. Urinary creatinine and albumin were measured using the Cobas Integra 400/800 system (Roche Diagnostics, Indianopolis, IN). At the first visit, Sickle cell trait (SCT) was determined in Black participants by genotyping using a TaqMan SNP Genotyping Assay (Applied Biosystems, Waltham, MA; ThermoFisher Scientific, Waltham, MA).^18^

C-reactive protein (CRP) was measured using a high-sensitivity, particle-enhanced immunonepholometric assay on the BNII nephelometer (N High Sensitivity CRP, Dade Behring Inc, Deerfield, IL now Siemens AG, Marburg, Germany). Hemoglobin was measured as a component of a hemogram, performed by automated cell counting on a Beckman Coulter LH 755 Hematology Workcell (Beckman Coulter, Incorporated, Fullerton, CA). Hemogram measurements began after 1/3 of the participants were recruited and examined. Assay methods for CRP and hemoglobin were the same at both visits.

### Inclusion of participants

From the initial 30,239 participants, HbA1c was measured at the first visit in a random sample of 3,276 participants, stratified on gender, race and age. HbA1c was measured at the second visit in 1,246 participants of this stratified random sample. After exclusion of participants with missing diabetes status or with diabetes at either assessment, we studied 917 participants without diabetes at both visits (figure 1).

**Figure 1.**
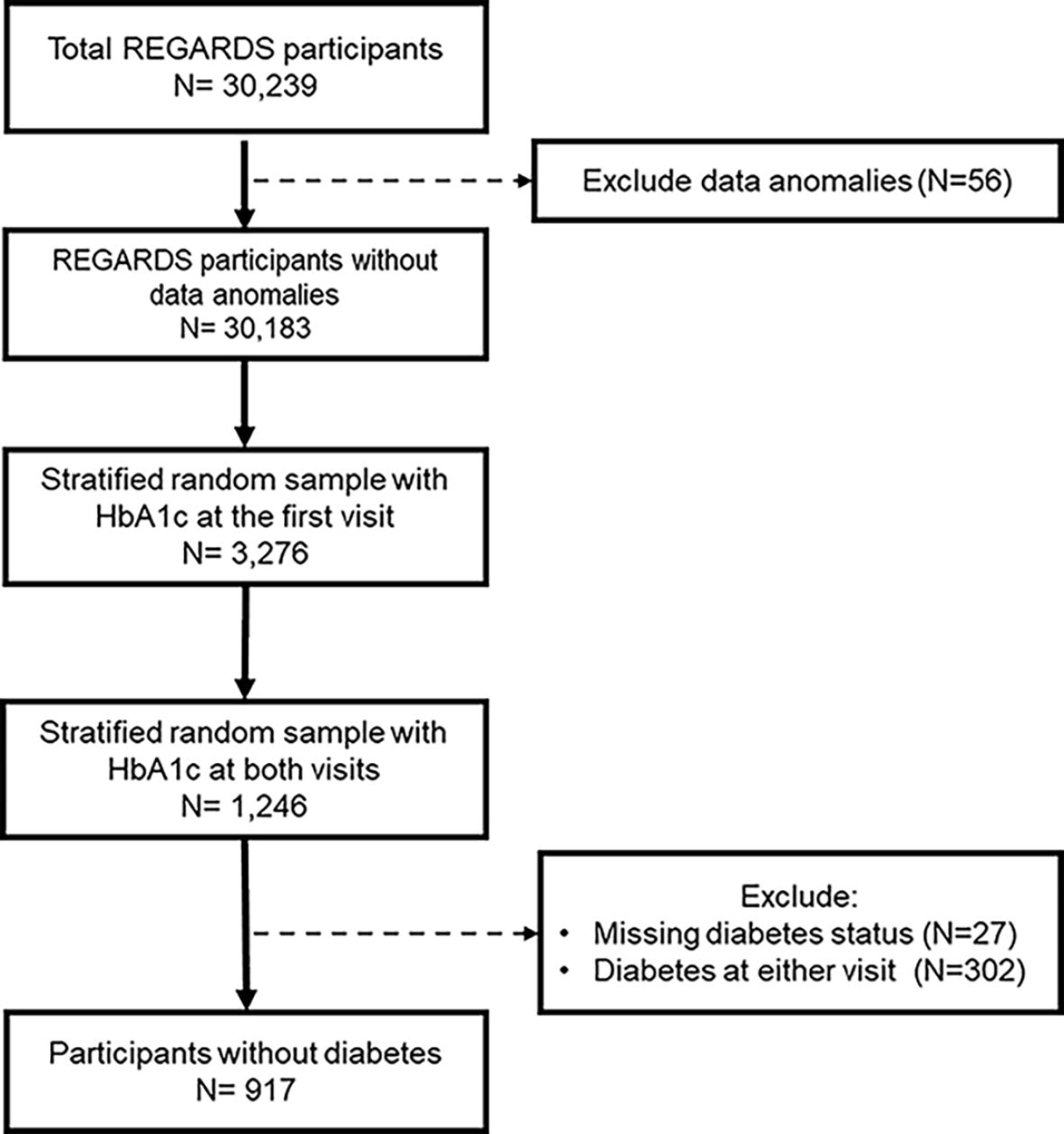
Flow diagram showing which study participants were included in the HbA1c analysis. From the initial 30,239 participants, 917 REGARDS participants without diabetes at Visit 1 (2003-07) and Visit 2 (2013-17) were included in the final analytic sample. Diabetes status was defined as fasting glucose ≥126 mg/dL or random glucose ≥200 mg/dL or self-reported diabetes or use of oral medications or insulin.

### Statistical methods

HbA1c data from the pilot study was plotted in scatterplots. Pearson correlations were calculated for comparison of each HbA1c assay method.

Sociodemographic, lifestyle, and laboratory characteristics were tabulated at both REGARDS visits. At each visit, linear regression was used to relate fasting plasma glucose and other clinical factors to HbA1c level in age, race, and gender-adjusted models. For skewed continuous variables (ACR, triglycerides and CRP), natural log transformation was applied.

The percentage of participants classified as having diabetes based only on HbA1c ≥6.5% at each visit was tabulated.

All statistical tests were two-sided, and p-values were considered statistically significant at alpha <0.05. Analyses were conducted using SAS version 9.4 (SAS Institute, Cary, NC, USA) and R version 4.1.0.

## RESULTS

In the pilot study (figure 2a-c), HbA1c measurements for whole blood samples (fresh or frozen) strongly correlated with measurements for packed cell samples (r ≥0.84).

**Figure 2.**
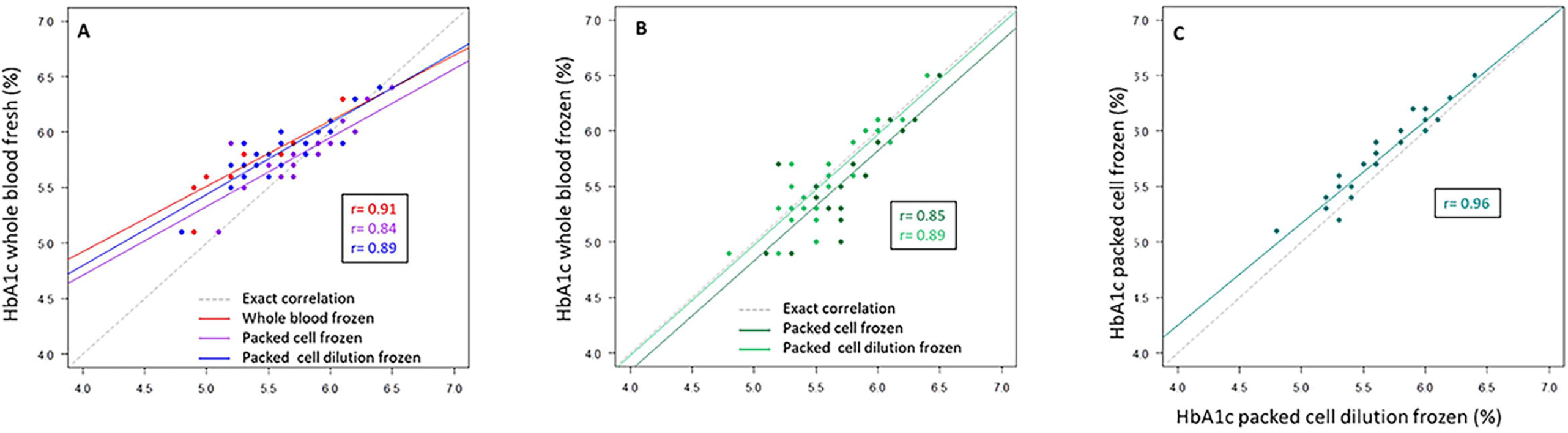
Pilot study (N=27) comparing hemoglobin A1c (HbA1c) in whole blood and packed cell samples: (A) Correlations between HbA1c measurements from fresh whole blood samples and other methods (frozen whole blood, frozen packed cells, and diluted frozen packed cells); (B) Correlations between HbA1c measurements from frozen whole blood samples and packed cells methods; (C) Correlations between HbA1c measurements from neat and diluted packed cells.

Among the studied REGARDS participants without diabetes at either visit, Table 1 shows that 40.7% were Black, and 44.9% had a college education or higher. About half were women, never-smokers and had baseline income ≥$35K. The mean fasting glucose and HbA1c at visit 1 and visit 2 were similar; glucose 91 and 90 mg/dL respectively, and HbA1c 5.8% and 5.7%. Lipid measures improved over time, and blood pressure, CRP and hemoglobin were slightly lower at the second than first visit. Kidney function moderately decreased over time while BMI remained stable.

**Table 1.** Baseline characteristics of 917 REGARDS participants without diabetes over 10 years (2003-2016)

| Characteristic | Visit |  |  |  |
| --- | --- | --- | --- | --- |
|  | Visit 1 | N | Visit 2 | N |
| Mean (SD), median (IQR)* or % |  |  |  |  |
| **Age years** | 63.3 (11.1) | 917 | 72.5 (11.2) | 917 |
| **Baseline gender** |  | 917 |  |  |
| Women | 52.8 |  |  |  |
| **Baseline race** |  | 917 |  |  |
| Black | 40.7 |  |  |  |
| **Baseline education** |  | 917 |  |  |
| Less than high school | 7.8 |  |  |  |
| High school graduate | 22.2 |  |  |  |
| Some college | 25.0 |  |  |  |
| College graduate and above | 44.9 |  |  |  |
| **Baseline income** |  | 917 |  |  |
| less than \$20k | 11.8 | | | |
| \$20k-\$34k | 22.0 | | | |
| \$35k-\$74k | 30.4 | | | |
| \$75k and above | 24.2 | | | |
| Refused | 11.6 |  |  |  |
| **Smoking** |  | 913 |  | 896 |
| Never | 50.5 |  | 50.1 |  |
| Past | 38.4 |  | 39.9 |  |
| Current | 10.7 |  | 7.7 |  |
| **Systolic blood pressure mmHg** | 125 (16) | 914 | 124 (15) | 917 |
| **Diastolic blood pressure mmHg)** | 76 (9) | 914 | 73 (9) | 917 |
| **BMI kg/m<sup>2</sup>** | 28.1 (5.3) | 911 | 28.1 (6.0) | 912 |
| **Sickle Cell Trait<sup>#</sup>** | 8.1 | 344 |  |  |
| **HbA1c %** | 5.8 (0.4) | 917 | 5.7 (0.4) | 917 |
| **Fasting Glucose mg/dL<sup>§</sup>** | 91 (9) | 811 | 90 (12) | 778 |
| **LDL mg/dL** | 116 (33) | 902 | 104 (35) | 915 |
| **HDL mg/dL** | 53 (16) | 912 | 58 (18) | 917 |
| **Triglycerides mg/dL<sup>\*</sup>** | 102 [76-138] | 916 | 96 [72-130] | 917 |
| **ACR mg/g<sup>\*</sup>** | 6.3 [4.0-11.0] | 892 | 9.2 [5.1-18.3] | 859 |
| **Serum Creatinine mg/dL** | 0.9 (0.2) | 917 | 1.0 (0.4) | 917 |
| **Estimated GFR mL/min/1.73 m<sup>2</sup>** | 88 (18) | 917 | 73 (21) | 917 |
| **CRP mg/L<sup>\*</sup>** | 1.8 [0.8-4.0] | 903 | 1.6 [0.8-4.1] | 894 |
| **Hemoglobin g/dL<sup>†</sup>** | 13.8 (1.4) | 665 | 13.5 (1.5) | 848 |

LDL: low-density lipoprotein; HDL: high-density lipoprotein; ACR: albumin-to-creatinine ratio; GFR: glomerular filtration rate; SBP: systolic blood pressure; DBP: diastolic blood pressure; CRP: C-reactive protein; BMI: body mass index; SCT: sickle cell trait; SD: standard deviation; IQR: interquartile range
\*: median (IQR) shown
#: Sickle cell trait genotyping was done for Black participants only.
§: participants not fasting were not included in analysis of glucose correlation with HbA1c
†: Hemoglobin measurements were not collected before May 2004<sup>36</sup>

Table 2 shows that associations between concurrent fasting glucose and HbA1c were similar using the two different methods for HbA1c determination, i.e., at both visits. Each 10mg/dL increment of glucose was associated with 0.08% (95%CI: 0.05-0.11%) higher HbA1c using frozen packed cells from the first visit and 0.10% (95%CI: 0.08-0.12%) higher HbA1c using the frozen cell layer from the second visit. Likewise, weak associations of HbA1c with age, gender, systolic blood pressure, BMI, HDL, triglycerides, CRP, and hemoglobin were similar at both visits. A stronger association between race and HbA1c was observed at the first visit compared to the second visit. Compared to White race, Black race was associated with 0.33% (95% CI=0.28-0.38%) higher HbA1c at the first visit and 0.15% (95% CI=0.10-0.19%) higher HbA1c at the second visit. both visits. Other variables in Table 2 were not associated with HbA1c at either visit.

**Table 2.** Association of Hemoglobin A1c with study variables of interest in 917 REGARDS participants without diabetes at Visit 1 (2003-07) and Visit 2 (2013-16)

| Characteristic | Difference in HbA1C by each Characteristic; $\beta$ ( 95% CI)* | |
| --- | --- | --- |
|  | Visit 1 (Frozen Packed Cells) | Visit 2 (Frozen Cell Layer) |
| **Similar Positive Association in Both Sample Types** |  |  |
| Fasting Glucose (per 10 mg/dL) | 0.08 (0.05, 0.10) | 0.11 (0.09, 0.13) |
| **Similar Weak Association in Both Sample Types** |  |  |
| Age (per 10 years) | 0.06 (0.04, 0.08) | 0.02 ( -0.001, 0.04) |
| Gender (Female) | 0.08 (0.04, 0.13) | 0.04 ( -0.01, 0.09) |
| Systolic blood pressure (per 10mmHg) | 0.03 (0.01, 0.04) | 0.02 ( -0.0005, 0.03) |
| BMI (per 5kg/m<sup>2</sup>) | 0.04 (0.02, 0.07) | 0.03 (0.01, 0.05) |
| HDL (per 5 mg/dL) | -0.01 ( -0.02, -0.005) | -0.02 ( -0.02, -0.008) |
| Log Triglycerides<sup>#</sup> (per 1 SD) | 0.03 (0.001, 0.05) | 0.05 (0.02, 0.07) |
| Log CRP<sup>#</sup> (per 1SD) | 0.04 (0.02, 0.07) | 0.05 (0.03, 0.08) |
| Hemoglobin (per 1g/dL) | -0.03 ( -0.06, -0.007) | -0.02 ( -0.04, 0.004) |
| **Different Association at Second Visit** |  |  |
| Race (Black) | 0.33 (0.28, 0.38) | 0.15 (0.10, 0.19) |
| **No Association at Either Visit** |  |  |
| Diastolic blood pressure (per 5mmHg) | 0.009 ( -0.02, 0.04) | -0.003 ( -0.03, 0.02) |
| LDL (per 10 mg/dL) | 0.003 ( -0.005, 0.01) | 0.002 ( -0.005, 0.009) |
| Log ACR<sup>#</sup> (1 per SD) | 0.02 ( -0.003, 0.05) | 0.003 ( -0.02, 0.03) |
| Serum Creatinine (per 0.2 mg/dL) | 0.02 ( -0.005, 0.05) | 0.004 ( -0.01, 0.019) |
| Estimated GFR (per 10 mL/min/1.73 m<sup>2</sup>) | -0.01 ( -0.03, 0.003) | -0.003 ( -0.02, 0.01) |
| Sickle cell trait (yes)<sup>§</sup> | -0.21 ( -0.59, 0.17)<sup>†</sup> | -0.06 ( -0.51, 0.39)<sup>†</sup> |

\*: Age, gender and race-adjusted
#: Standardized
§: Sickle cell trait genotyping was done for Black participants only.
†: Sickle cell trait was adjusted for age and gender, but not adjusted for race
BMI: body mass index; HDL: high-density lipoprotein; CRP: C-reactive protein; LDL: low-density lipoprotein; ACR: albumin-to-creatinine ratio; GFR: glomerular filtration rate; SD: standard deviation

Among these participants classified as not meeting diabetes criteria using fasting glucose or clinical history, using the HbA1c diagnostic cut-off of 6.5%, 3.5% were classified as meeting diabetes cut-point at the first visit and 2.3 % at the second visit (table 3).

**Table 3.**
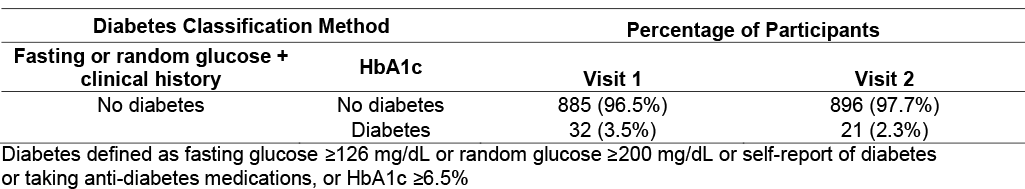
Classification of diabetes based on HbA1c at two REGARDS Visits. HbA1c was measured at Visit 1 (2003-07) in frozen packed cells, and at Visit 2 (2013-16) in frozen whole blood.

| Diabetes Classification Method |  | Percentage of Participants |  |
| --- | --- | --- | --- |
| Fasting or random glucose + clinical history | HbA1c | Visit 1 | Visit 2 |
| No diabetes | No diabetes | 885 (96.5%) | 896 (97.7%) |
|  | Diabetes | 32 (3.5%) | 21 (2.3%) |

Diabetes defined as fasting glucose $\geq 126$ mg/dL or random glucose $\geq 200$ mg/dL or self-report of diabetes or taking anti-diabetes medications, or HbA1c $\geq 6.5\%$

## DISCUSSION

This study investigated reliability of REGARDS methods to measure HbA1c in frozen stored samples for epidemiology research. In a pilot study, different measurement methods were highly correlated. HbA1c values from frozen packed cells stored at -80°C for up to 14 years correlated in the expected manner with fasting glucose values and several known correlates of HbA1c. Further, frozen whole blood results from samples at a second visit correlated similarly as the baseline samples with glucose and other correlates. There was excellent agreement between HbA1c and glucose criteria for defining absence of diabetes.

Pre-analytical factors including sample freezing and storage time affect use of HbA1c values in clinical practice. There is limited published data on HbA1c testing of samples frozen for many years in the context of epidemiologic or other research studies. Selvin et al reported strong correlation between HbA1c results obtained from frozen whole blood samples before and after 11-14 years of storage at -70°C in the Atherosclerosis Risk in Communities Study.^6^ Testing at both timepoints was performed using ion-exchange HPLC assays, and while correlation between short-term and long-term stored sample results was high (r = 0.97), an upward bias of 0.35 percentage points was observed in long-term stored samples. It has been noted that hemoglobin adducts formed as a result of degradation can influence the charge characteristics of the molecule and interfere with ion-exchange HPLC assays,^7^ but whether this underlies the observed bias in the previously referenced study is unclear. The present study used an immunoassay, which should not be affected by alterations in charge. In a published comparison of effects of whole blood storage on HbA1c measurements using various types of assays (boronate-affinity HPLC, ion-exchange HPLC, and immunoassay), the immunoassay exhibited the best stability in samples stored at -20°C.^8^ Our pilot data also showed sample types should not affect analyses. In our pilot study, HbA1c values for frozen or fresh whole blood samples strongly correlated with HbA1c values for frozen packed cell samples (r≥0.84). In REGARDS, HbaA1c values and correlates were similar from the first to the second visit. In agreement, in previous research, among people without diabetes over time, HbA1c changed minimally over a five-year period.^19^ This supports the validity of methods for HbA1c measurement used in our study.

Consistent with our findings, a previous study in participants of The Malaysian Cohort study found 3.3% of individuals with fasting glucose <126 mg/dL were not classified as meeting the diabetes cut-point using HbA1c ≥6.5%. ^20^ Among people without diabetes, HbA1c is associated with fasting plasma glucose,^21^ age,^21-23^ gender,^24^ race,^25^ systolic blood pressure,^26^ BMI,^21,27^ HDL,^27^ triglycerides,^27^ CRP,^23^ hemoglobin, ^21^ and kidney function.^28,29^ Most of those associations were similarly modest compared to our findings. In the present study, associations of HbA1c with fasting plasma glucose, triglycerides, HDL, BMI, and CRP were of similar magnitude at both visits. These findings provide support for our measurement method validity. We saw a stronger association between race and HbA1c at the first visit compared to the second visit, although the overall association was small. The difference over time could be because packed cells were used at the first visit while frozen whole blood samples were used at the second visit, or due to differences in health status over time not accounted for. Although the pilot study indicated a strong correlation in HbA1c values between frozen whole blood samples and packed cells, it is still possible that the difference in sample types affected this association.

In concordance with previous findings,^26,30^ this study showed HbA1c was not associated with diastolic blood pressure and LDL in people without diabetes. Our results also showed that HbA1c was not associated with sickle cell trait in Black people. A previous study, which used HPLC to measure HbA1c, reported that HbA1c values were lower in Black Americans with sickle cell trait compared to those without.^31^ Measurement of HbA1c depends on a normal erythrocyte life span^32^, and hemoglobinopathies have varying effects on HbA1c concentrations.^32,33^ Unlike people homozygous for hemoglobin S variant (HbS), people with sickle cell trait may have normal erythrocyte survival, and HbA1c can be used to assess their glycemic status as long as HbS does not interfere with the assay method or glucose binding.^32^ HbS does not interfere with the immunoassay method used in our study,^34^ but it affects HPLC methods.^32^

HbA1c was not associated with kidney function. This conflicts with prior evidence that showed associations.^28,29^ This difference in findings could be due to different measurement methods for HbA1c (HPLC vs. immunoassay), or that the weak prior observed associations were observable as these studies had a very large sample size.

There are limitations to consider in this research. To achieve our goal of assessing validity of our measurement methods, the studied sample only included people without diabetes, so findings do not apply to those with diabetes. Prior to storage, blood samples were frozen on ice and shipped overnight. A recent report indicated that sample transport could cause hemolysis and affect HbA1c levels.^35^ We did not measure HbA1c in fresh whole blood compared to measurements years later in frozen samples, but instead presented the pilot study, correlates of HbA1c in stored samples at two time points as a way to indirectly assess reliability.

Notwithstanding these limitations, there are several strengths. Our study spanned a considerable amount of time and included measures of HbA1c in both whole blood samples and packed cells. We also investigated several known correlates of HbA1c. To our knowledge, this is one of the few longitudinal studies to investigate HbA1c over a long period of time in people without diabetes and to examine HbA1c in samples after long-term storage and in whole blood samples and packed cells.

In conclusion, in REGARDS, HbA1c measurements appeared valid for both frozen whole blood and frozen packed cells samples stored at -80°C for up to 14 years in people without diabetes. Future studies should extend these findings to people with diabetes and address calibration of HbA1c to detect diabetes. Future studies could also determine whether HbA1c measured with frozen packed cells or whole blood stored for years predicts incident diabetes and cardiovascular outcomes.

## Supporting information

Abstract

## Data Availability

Data are available from the University of Alabama at Birmingham. Requests to access these datasets should be directed to.

## ACKNOWLEDGMENT

This research project is supported by cooperative agreement U01 NS041588 co-funded by the National Institute of Neurological Disorders and Stroke (NINDS) and the National Institute on Aging (NIA), National Institutes of Health, Department of Health and Human Service. The content is solely the responsibility of the authors and does not necessarily represent the official views of the NINDS or the NIA. Representatives of the NINDS were involved in the review of the manuscript but were not directly involved in the collection, management, analysis or interpretation of the data. Additional support was from the NIH National Institute of General Medical Sciences P20GM135007 and the NIH Office of the Director UG3OD023316. The authors thank the other investigators, the staff, and the participants of the REGARDS study for their valuable contributions. A full list of participating REGARDS investigators and institutions can be found at http://www.regardsstudy.org.

## DISCLAIMER

This article was prepared while Stephanie Tison was employed at the University of Alabama at Birmingham as a Graduate Research Assistant for the REGARDS research project. The opinions expressed in this article are the author’s own and do not reflect the view of the National Institutes of Health, the Department of Health and Human Services, or the United States government.

## References

1. Group UPDSU. Intensive blood-glucose control with sulphonylureas or insulin compared with conventional treatment and risk of complications in patients with type 2 diabetes (UKPDS 33). UK Prospective Diabetes Study (UKPDS) Group. Lancet (London, England). 1998;352:837–853.

2. Association AD. Diagnosis and classification of diabetes mellitus. Diabetes care. 2010;33 Suppl 1:S62–69. doi: 10.2337/dc10-S062

3. Youngman LD, Clark S, Manley S, Peto R, Collins R. Reliable measurement of glycated hemoglobin in frozen blood samples: implications for epidemiologic studies. Clin Chem. 2002;48:1627–1629.

4. Rolandsson O, Marklund SL, Norberg M, Agren A, Hagg E. Hemoglobin A1c can be analyzed in blood kept frozen at -80 degrees C and is not commonly affected by hemolysis in the general population. Metabolism. 2004;53:1496–1499. doi: 10.1016/j.metabol.2004.04.015

5. Jones W, Scott J, Leary S, Stratton F, Smith S, Jones R, Day A, Ness A. Stability of whole blood at -70 degrees C for measurement of hemoglobin A(1c) in healthy individuals. Clin Chem. 2004;50:2460–2461. doi: 10.1373/clinchem.2004.038521

6. Selvin E, Coresh J, Jordahl J, Boland L, Steffes MW. Stability of haemoglobin A1c (HbA1c) measurements from frozen whole blood samples stored for over a decade. Diabetic medicine : a journal of the British Diabetic Association. 2005;22:1726–1730. doi: 10.1111/j.1464-5491.2005.01705.x

7. Little RR, Rohlfing CL, Tennill AL, Connolly S, Hanson S. Effects of sample storage conditions on glycated hemoglobin measurement: evaluation of five different high performance liquid chromatography methods. Diabetes technology & therapeutics. 2007;9:36–42. doi: 10.1089/dia.2006.0055

8. Rohlfing CL, Hanson S, Tennill AL, Little RR. Effects of whole blood storage on hemoglobin a1c measurements with five current assay methods. Diabetes technology & therapeutics. 2012;14:271–275. doi: 10.1089/dia.2011.0136

9. Liotta L, Di Franco A, Pazzagli M, Luconi M. Glycated hemoglobin (HbA1c) measurement in frozen whole blood depends on baseline values of fresh samples. Analytical and bioanalytical chemistry. 2013;405:429–434. doi: 10.1007/s00216-012-6480-y

10. Bergmann K, Sypniewska G. The influence of sample freezing at - 80 degrees C for 2-12 weeks on glycated haemoglobin (HbA1c) concentration assayed by HPLC method on Bio-Rad D-10((R)) auto analyzer. Biochem Med (Zagreb). 2016;26:346–352. doi: 10.11613/bm.2016.038

11. Howard VJ, Cushman M, Pulley L, Gomez CR, Go RC, Prineas RJ, Graham A, Moy CS, Howard G. The reasons for geographic and racial differences in stroke study: objectives and design. Neuroepidemiology. 2005;25:135–143. doi: 10.1159/000086678

12. Rhea JM, Molinaro R. Pathology consultation on HbA(1c) methods and interferences. Am J Clin Pathol. 2014;141:5–16. doi: 10.1309/ajcpq23gttmlaevl

13. Gillett SR, Boyle RH, Zakai NA, McClure LA, Jenny NS, Cushman M. Validating laboratory results in a national observational cohort study without field centers: the Reasons for Geographic and Racial Differences in Stroke cohort. Clin Biochem. 2014;47:243–246. doi: 10.1016/j.clinbiochem.2014.08.003

14. Howard G, Cushman M, Kissela BM, Kleindorfer DO, McClure LA, Safford MM, Rhodes JD, Soliman EZ, Moy CS, Judd SE, et al. Traditional risk factors as the underlying cause of racial disparities in stroke: lessons from the half-full (empty?) glass. Stroke. 2011;42:3369–3375. doi: 10.1161/STROKEAHA.111.625277

15. Friedewald WT, Levy RI, Fredrickson DS. Estimation of the concentration of low-density lipoprotein cholesterol in plasma, without use of the preparative ultracentrifuge. Clin Chem. 1972;18:499–502.

16. Kurella Tamura M, Wadley V, Yaffe K, McClure LA, Howard G, Go R, Allman RM, Warnock DG, McClellan W. Kidney function and cognitive impairment in US adults: the Reasons for Geographic and Racial Differences in Stroke (REGARDS) Study. Am J Kidney Dis. 2008;52:227–234. doi: 10.1053/j.ajkd.2008.05.004

17. Levey AS, Stevens LA, Schmid CH, Zhang YL, Castro AF, 3rd, Feldman HI, Kusek JW, Eggers P, Van Lente F, Greene T, et al. A new equation to estimate glomerular filtration rate. Ann Intern Med. 2009;150:604–612. doi: 10.7326/0003-4819-150-9-200905050-00006

18. Cahill CR, Leach JM, McClure LA, Irvin MR, Zakai NA, Naik R, Unverzagt F, Wadley VG, Hyacinth HI, Manly J, et al. Sickle cell trait and risk of cognitive impairment in African-Americans: The REGARDS cohort. EClinicalMedicine. 2019;11:27–33. doi: 10.1016/j.eclinm.2019.05.003

19. Halalau A, Roy S, Hegde A, Khanal S, Langnas E, Raja M, Homayouni R. Risk factors associated with glycated hemoglobin A1c trajectories progressing to type 2 diabetes. Ann Med. 2023;55:371–378. doi: 10.1080/07853890.2022.2164347

20. Abdul Murad NA, Abdullah N, Kamaruddin MA, Abd Jalal N, Ismail N, Yusof NAM, Mustafa N, Jamal R. Discordance between Fasting Plasma Glucose (FPG) and HbA1c in Diagnosing Diabetes and Pre-diabetes in The Malaysian Cohort. J ASEAN Fed Endocr Soc. 2021;36:127–132. doi: 10.15605/jafes.036.02.02

21. Jansen H, Stolk RP, Nolte IM, Kema IP, Wolffenbuttel BH, Snieder H. Determinants of HbA1c in nondiabetic Dutch adults: genetic loci and clinical and lifestyle parameters, and their interactions in the Lifelines Cohort Study. J Intern Med. 2013;273:283–293. doi: 10.1111/joim.12010

22. Pani LN, Korenda L, Meigs JB, Driver C, Chamany S, Fox CS, Sullivan L, D’Agostino RB, Nathan DM. Effect of aging on A1C levels in individuals without diabetes: evidence from the Framingham Offspring Study and the National Health and Nutrition Examination Survey 2001-2004. Diabetes care. 2008;31:1991–1996. doi: 10.2337/dc08-0577

23. Wu T, Dorn JP, Donahue RP, Sempos CT, Trevisan M. Associations of serum C-reactive protein with fasting insulin, glucose, and glycosylated hemoglobin: the Third National Health and Nutrition Examination Survey, 1988-1994. Am J Epidemiol. 2002;155:65–71. doi: 10.1093/aje/155.1.65

24. Ma Q, Liu H, Xiang G, Shan W, Xing W. Association between glycated hemoglobin A1c levels with age and gender in Chinese adults with no prior diagnosis of diabetes mellitus. Biomed Rep. 2016;4:737–740. doi: 10.3892/br.2016.643

25. Cavagnolli G, Pimentel AL, Freitas PA, Gross JL, Camargo JL. Effect of ethnicity on HbA1c levels in individuals without diabetes: Systematic review and meta-analysis. PLoS One. 2017;12:e0171315. doi: 10.1371/journal.pone.0171315

26. Liu L, Zhen D, Fu S, Sun W, Li H, Zhao N, Hou L, Tang X. Associations of the baseline level and change in glycosylated hemoglobin A1c with incident hypertension in non-diabetic individuals: a 3-year cohort study. Diabetol Metab Syndr. 2022;14:54. doi: 10.1186/s13098-022-00827-8

27. Pai JK, Cahill LE, Hu FB, Rexrode KM, Manson JE, Rimm EB. Hemoglobin a1c is associated with increased risk of incident coronary heart disease among apparently healthy, nondiabetic men and women. J Am Heart Assoc. 2013;2:e000077. doi: 10.1161/JAHA.112.000077

28. Kang SH, Jung DJ, Choi EW, Cho KH, Park JW, Do JY. HbA1c Levels Are Associated with Chronic Kidney Disease in a Non-Diabetic Adult Population: A Nationwide Survey (KNHANES 2011-2013). PLoS One. 2015;10:e0145827. doi: 10.1371/journal.pone.0145827

29. Kang SH, Park JW, Do JY, Cho KH. Glycated hemoglobin A1c level is associated with high urinary albumin/creatinine ratio in non-diabetic adult population. Ann Med. 2016;48:477–484. doi: 10.1080/07853890.2016.1197412

30. Spessatto D, Brum L, Camargo JL. Oxidized LDL but not total LDL is associated with HbA1c in individuals without diabetes. Clin Chim Acta. 2017;471:171–176. doi: 10.1016/j.cca.2017.06.004

31. Lacy ME, Wellenius GA, Sumner AE, Correa A, Carnethon MR, Liem RI, Wilson JG, Sacks DB, Jacobs DR, Jr., Carson AP, et al. Association of Sickle Cell Trait With Hemoglobin A1c in African Americans. JAMA. 2017;317:507–515. doi: 10.1001/jama.2016.21035

32. Little RR, Roberts WL. A review of variant hemoglobins interfering with hemoglobin A1c measurement. J Diabetes Sci Technol. 2009;3:446–451. doi: 10.1177/193229680900300307

33. Liddy AM, Grundy S, Sreenan S, Tormey W. Impact of haemoglobin variants on the use of haemoglobin A1c for the diagnosis and monitoring of diabetes: a contextualised review. Ir J Med Sci. 2022. doi: 10.1007/s11845-022-02967-2

34. U.S. Department Of Health & Human Services Food and Drug Administration. cobas c 513 Analyzer, cobas c 513 Tina-quant HbA1cDx Gen.3 Assay approval letter K160571 510(k) Premarket Notification. 2016.

35. Koga M, Okumiya T, Ishibashi M. Sample transport and/or storage can cause falsely low HbA1c levels in blood cells measured by enzymatic assay. Diabetol Int. 2020;11:155–157. doi: 10.1007/s13340-019-00416-7

36. Panwar B, Judd SE, Warnock DG, McClellan WM, Booth JN, 3rd, Muntner P, Gutierrez OM. Hemoglobin Concentration and Risk of Incident Stroke in Community-Living Adults. Stroke. 2016;47:2017–2024. doi: 10.1161/STROKEAHA.116.013077

