## Supplementary material for "Measuring Hemoglobin A1C with Frozen Packed Cell and Frozen Whole Blood Samples in an Epidemiologic Study: The Reasons for Geographic and Racial Differences in Stroke (REGARDS) Study": Abstract

**GRAPHICAL ABSTRACT**

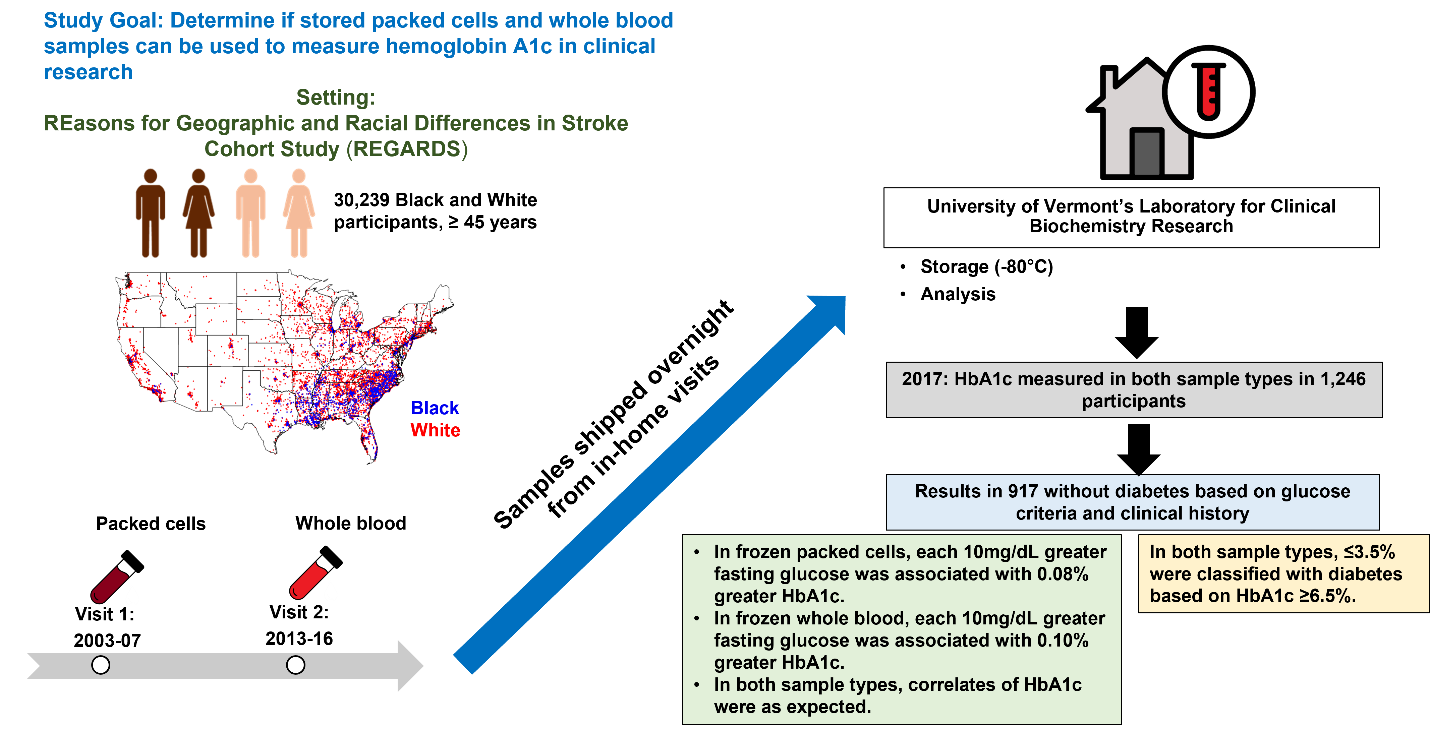

**Highlights**

- In REGARDS participants without diabetes, HbA1c values from frozen whole blood and packed cell samples under long-term storage correlated in the expected manner with fasting glucose values and several known correlates of HbA1c.
- These findings indicate that frozen whole blood and frozen packed cells stored over a long period of time are acceptable for study of the epidemiology of HbA1c.
